# Timely Epidemic Monitoring in the Presence of Reporting Delays: Anticipating the COVID-19 Surge in New York City, September 2020

**DOI:** 10.1101/2020.08.02.20159418

**Authors:** Jeffrey E. Harris

## Abstract

During a fast-moving epidemic, timely monitoring of case counts and other key indicators of disease spread is critical to an effective public policy response. We describe a nonparametric statistical method, originally applied to the reporting of AIDS cases in the 1980s, to estimate the distribution of reporting delays of confirmed COVID-19 cases in New York City during the late summer and early fall of 2020. During August 15 - September 26, the estimated mean delay in reporting was 3.3 days, with 87 percent of cases reported by 5 days from diagnosis. Relying upon the estimated reporting-delay distribution, we projected COVID-19 incidence during the most recent three weeks as if each case had instead been reported on the same day that the underlying diagnostic test had been performed. Applying our delay-corrected estimates to case counts reported as of September 26, we projected a surge in new diagnoses that had already occurred but had yet to be reported. Our projections were consistent with counts of confirmed cases subsequently reported by November 7. The resulting estimate of recently diagnosed cases could have had an impact on timely policy decisions to tighten social distancing measures. While the recent advent of widespread rapid antigen testing has changed the diagnostic testing landscape considerably, delays in public reporting of SARS-CoV-2 case counts remain an important barrier to effective public health policy.

## Introduction

Timely surveillance of the incidence of new cases is essential for effective control during an ongoing epidemic. When infections are detected primarily through voluntary testing of symptomatic individuals, as has been the case with the COVID-19 epidemic in the United States, there will be three main sources of incomplete or delayed reporting of new cases. First, there will be *underreporting* of new cases, especially among asymptomatic or mildly infected individuals who do not seek testing. Second, there will be a *testing delay* between the actual date when an individual becomes infected and the date when that individual is ultimately tested. Third, unless test samples are rapidly processed, there will be a further *reporting delay* between the date of testing and the date the test results are communicated by the reporting entity. The present research addresses the latter source of delay.

A statistical method for nonparametric or semiparametric estimation of the distribution of reporting delays was previous investigated in connection with delays in reporting of newly diagnosed AIDS cases during the 1980s.^1^ The estimated distribution of delays allowed the analyst to predict the actual incidence of AIDS cases well before all cases were fully reported. That statistical method is adapted here to daily reports of newly diagnosed cases of COVID-19 by the New York City Department of Health and Mental Hygiene during the late summer and early fall of 2020, when there was heightened concern about the possible emergence of a new wave of infection in the city. Our objective is to determine whether this approach could have alerted public officials to the coming surge before it became apparent from other indicators.

## Data

All data were downloaded from the New York City health department repository.^2^ The data consisted of a series of daily updates of a file named *case-hosp-death*.*csv*. In this report, we relied solely on the first two variables in each updated file, labeled *DATE_OF_INTEREST* and *CASE_COUNT*, which we interpreted, respectively, as the date of diagnosis and the cumulative number of confirmed COVID-19 cases so far diagnosed by that date. We did not rely on data on hospitalizations or deaths in this study.

Fig. 1 displays the reported numbers of test-confirmed, daily COVID-19 cases from June 21 through September 26, 2020. The horizontal axis measures the date of diagnosis, that is, the date on which the confirming diagnostic test was performed, rather than the date on which the case was subsequently reported by the health department.

**Fig. 1.**
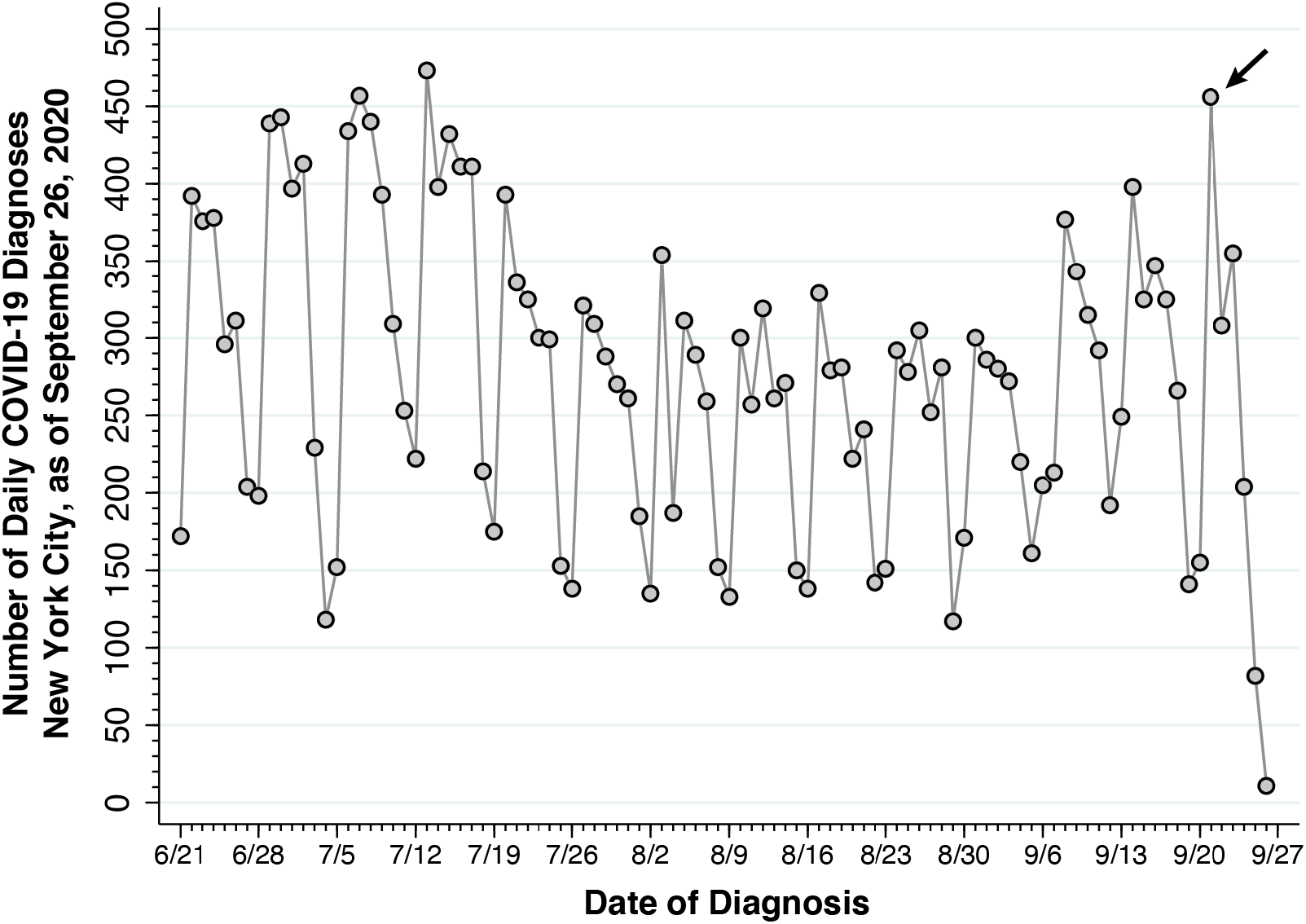
Confirmed Cases of COVID-19 by Date of Diagnosis, New York City, June 21 – September 26, 2020, Reported as of September 26, 2020. Reported cases rose to 456 on September 21 (arrow), nearly equaling the previous peak of 457 on July 7. Thereafter, during September 22 – 26, reported cases declined precipitously. As of September 26, only 11 cases were reported as diagnosed on that date.

During September 2020, there was increasing concern among public health officials that the low case counts reported during the summer were beginning to rise, and that the observed increase foreshadowed the onset of a new wave of SARS-CoV-2 infections in the city.^3^ During August, as Fig. 1 shows, daily reported case counts had settled down to between 100–350, depending on the day of the week. By September 26, as indicated by the arrow in the figure, number of cases diagnosed *and thus far reported* on September 21 had reached 456, nearly equaling the previous peak of 457 on July 7.

The precipitous decline in reported cases during September 22–26, however, posed a serious problem of data interpretation. It was widely acknowledged that recent case counts were significantly truncated as a result of reporting delays. Thus, the datapoint for September 26, showing only 11 cases, represented only those cases that were diagnosed *and* reported on that same date. In its website displaying trends in newly confirmed COVID-19 cases, the health department employed the usual workaround of advising website visitors that, as a result of delays in reporting, recent data were incomplete.

## Methods

### Statistical Analysis of Reporting Delays

From successive daily updates of the *case-hosp-death*.*csv* file, we computed the quantities *y*_*tu*_, corresponding to the number of confirmed infections diagnosed on date *t* but not reported until date *t* + *u*, that is, with a delay of *u* ≥ 0 days. For example, the version of the file *case-hosp-death*.*csv* showing all reports through 7/21/2020 indicated that 9 cases had been diagnosed on that date and thus far reported by that date. The following day’s version of *case-hosp-death*.*csv* indicated that a total of 65 cases had been diagnosed on 7/21/2020 and reported by 7/22/2020. Thus, we have *y*_*t*0_ = 9 and *y*_*t*1_ = 65 – 9 = 56, where *t* corresponds in this example to the diagnosis date 7/21/2020. The very next day’s version indicated that a total of 132 cases had been diagnosed on 7/21/2020 and reported by 7/23/2020. Thus, we have *y*_*t*2_ = 132 – 65 = 67. We used this method of successive differences to recover the underlying quantities {*y*_*tu*_}, which formed the basic data for our analysis.

Our statistical approach followed earlier work.^1^ Let the possible dates of diagnosis *t* range from 0 to *T*, where *T* > 0 is the last date on which we have received case reports, which we’ll call the *cutoff date*. Let the duration of reporting delay *u* range from 0 to *n*, where *n* > 0 is assumed to be the longest possible reporting delay. We further assume that *T* > *n* > 0. As a result of this assumption, our sample is bifurcated into two parts, which we call the *early* and *late* parts, respectively. The early part corresponds to dates of diagnosis *t* = 0, …, *T* − *n*. For these dates, we have by assumption a complete set {*y*_*tu*_, *u* = 0,1, …, *n*} of all reported cases diagnosed on each date. The late part corresponds to subsequent dates of diagnosis *t* = *T* − *n* + 1, …, *T*. For these dates, we have only a truncated set {*y*_*tu*_, *u* = 0,1,…,*T* − *t*} of reported cases diagnosed on each date, as some diagnoses have not yet been reported by the cutoff date *T*.

We considered the simplest model where the distribution of delays was independent of the date of diagnosis or any other observable, exogenous variable. That is, the probability that a case diagnosed at date *t* will be reported with delay *u* is *α*_*u*_, where 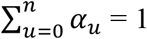. Let *α* = (*α*_0_, *α*_1_, …, *α*_*n*_) denote the vector all parameters *α*_*u*_. Extensions of this basic model, including a variation in which 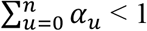, are described elsewhere.^1^

The basic idea is to estimate the delay distribution *α* from our observed data, and then use the estimate 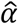 to project the total number of cases diagnosed on a given date, including diagnoses yet to be reported. In general, we define 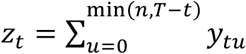 as the total number of cases diagnosed on date *t* that have so far been reported by the cutoff date *T*. In the early part of the sample, for any date of diagnosis *t* = 0, …, *T* − *n*, this marginal sum simplifies to 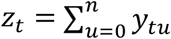 and represents the total number of cases diagnosed on that date. So, conditional on the marginal sums *z*_*t*_, we have the projected number of cases *ζ*(*α*) = *z*_*t*_, which is independent of *α*. Since we have already observed all the cases diagnosed on date *t*, there is nothing unknown to project.

In the late part of the sample, for any date of diagnosis *t* = *T* − *n* + 1, …, *T*, we can instead write the marginal sums as 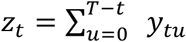. The projected number of cases diagnosed at date *t* will depend on the parameters as *ζ*_*t*_ (*α*) = *z*_*t*_/Ω_*t*_ (*α*), where 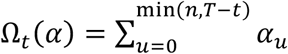 is the estimated probability that a case diagnosed at date *t* will be reported by the cutoff date *T*.

We assume that the counts {*y*_*tu*_} are the realizations of independent Poisson random variables. Given the marginal sums *z*_*t*_, the conditional likelihood of the parameters *α* is maximized by the following iterative procedure, which is equivalent to the EM algorithm.^4^ Let 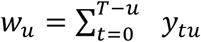 denote the total number of cases reported with a delay of *u* days, summed over all dates of diagnosis *t*. We start with initial estimates 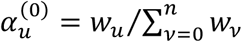 for all *u* = 0, …, *n*. At iteration *k* = 0,1,2, …, with provisional parameters *α*^(*k*)^, we update our parameters to 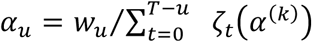, where the denominator is the projected total number of diagnosed cases for which a delay *u* has been observed. To complete the iteration, we normalize to get 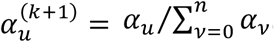. We continue to iterate until ⌈*α*^(*k*+1)^ − *α*^(*k*)^ ⌉ is arbitrarily small. Once we’ve converged on an estimate 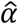, the projected case counts are 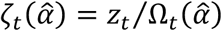 for all *t* = 0, … *T*.

## Results

### Distribution of Reporting Delays

We estimated the probability distribution *α* of reporting delays up to a maximum of *n* = 21 days from data {*y*_*tu*_} on case counts reported during August 15 – September 26, 2020. Thus, we took August 15 as the initial observation date *t* = 0, while September 26 was date *t* = *T* = 42. As a result, the early part of our sample, that is, the range of dates *t* for which the observed case counts {*y*_*tu*_, *u* = 0,…,21} were complete, ran from August 15 through September 5. The late part of our sample, in which the observations on *y*_*tu*_ were truncated, ran from September 16 – 26.

Fig.2 shows the estimated probability distribution 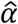 of reporting delays. Only 4.1 percent of confirmed COVID-19 cases were reported on the same day that the underlying diagnostic test was performed, that is, 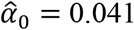. The estimated probability of reporting within 5 days of diagnosis was 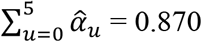. The mean reporting delay, conditional upon full reporting by *n* = 21 days, was 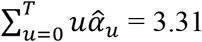 days.

**Fig. 2.**
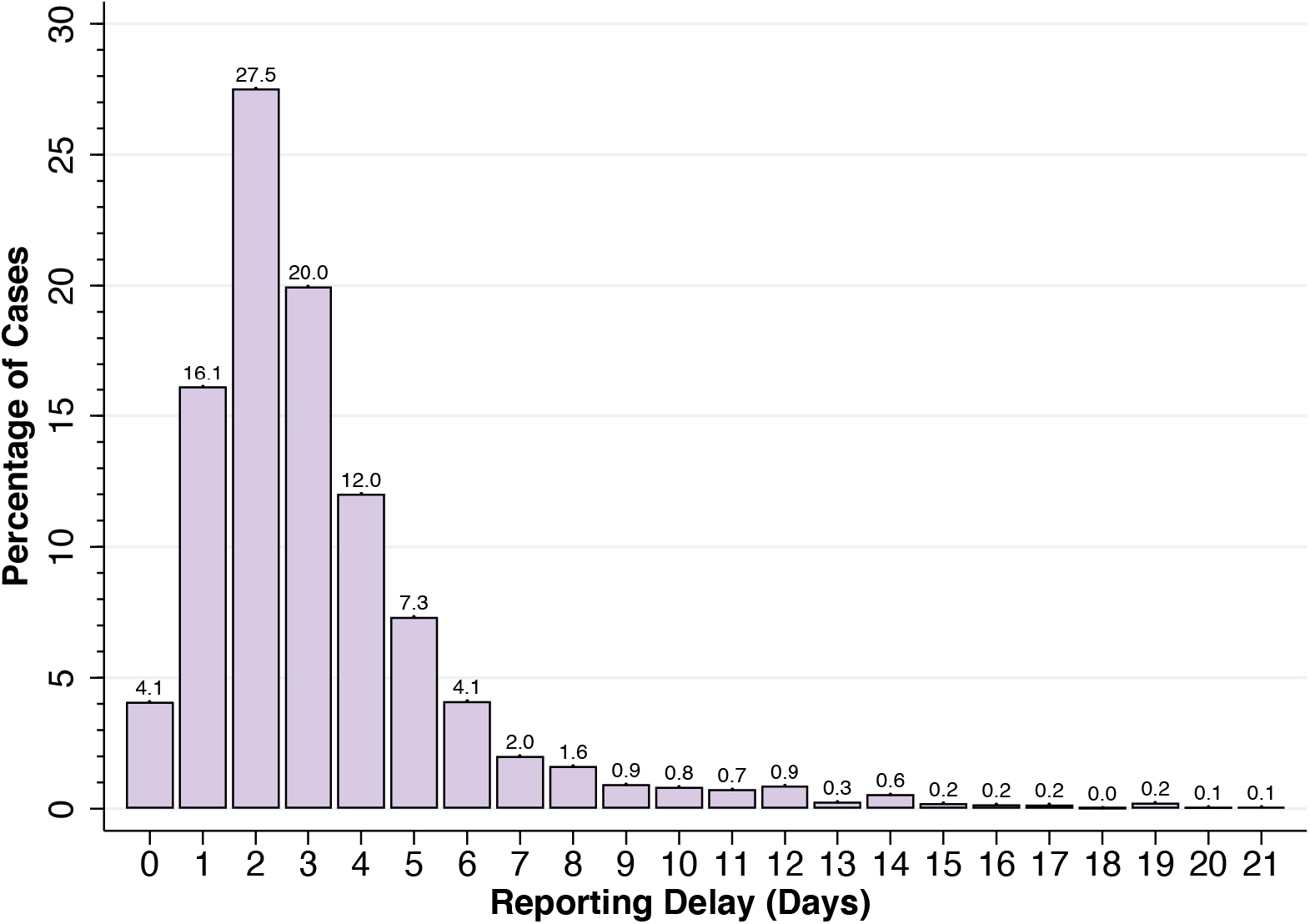
Estimated Distribution of Reporting Delays, Based Upon Confirmed COVID-19 Cases Reported During August 15 – September 26, 2020. The height of each bar (in percent) corresponds to 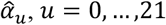. The mean delay, conditional upon full reporting by *n* = 21 days, was 3.31 days.

We observed shifts in the estimated distribution of reporting delays in New York City during the first six months of the COVID-19 epidemic. Reporting delays initially increased from the initial outbreak in early March through June 2020. From late June onward, however, the reporting delay distribution began to shift to the left, as shown by the comparison of the estimated cumulative distribution curves for the periods from June 21 – August 1 and from August 15 – September 26, shown in Appendix Fig. A. During the more recent interval from August 15 – September 26, however, the reporting delay distribution appeared to be stable.

### Incidence of COVID-19 Cases Corrected for Reporting Delays

The gray datapoints in Fig. 3 reproduce the observed counts of newly diagnosed COVID-19 cases during June 21 – September 26, 2020, as shown in Fig. 1 above. While only the counts *z*_*t*_ from August 15 (date *t* = 0) entered our estimation algorithm, we show the complete series back to June 21 to facilitate comparison.

**Fig. 3.**
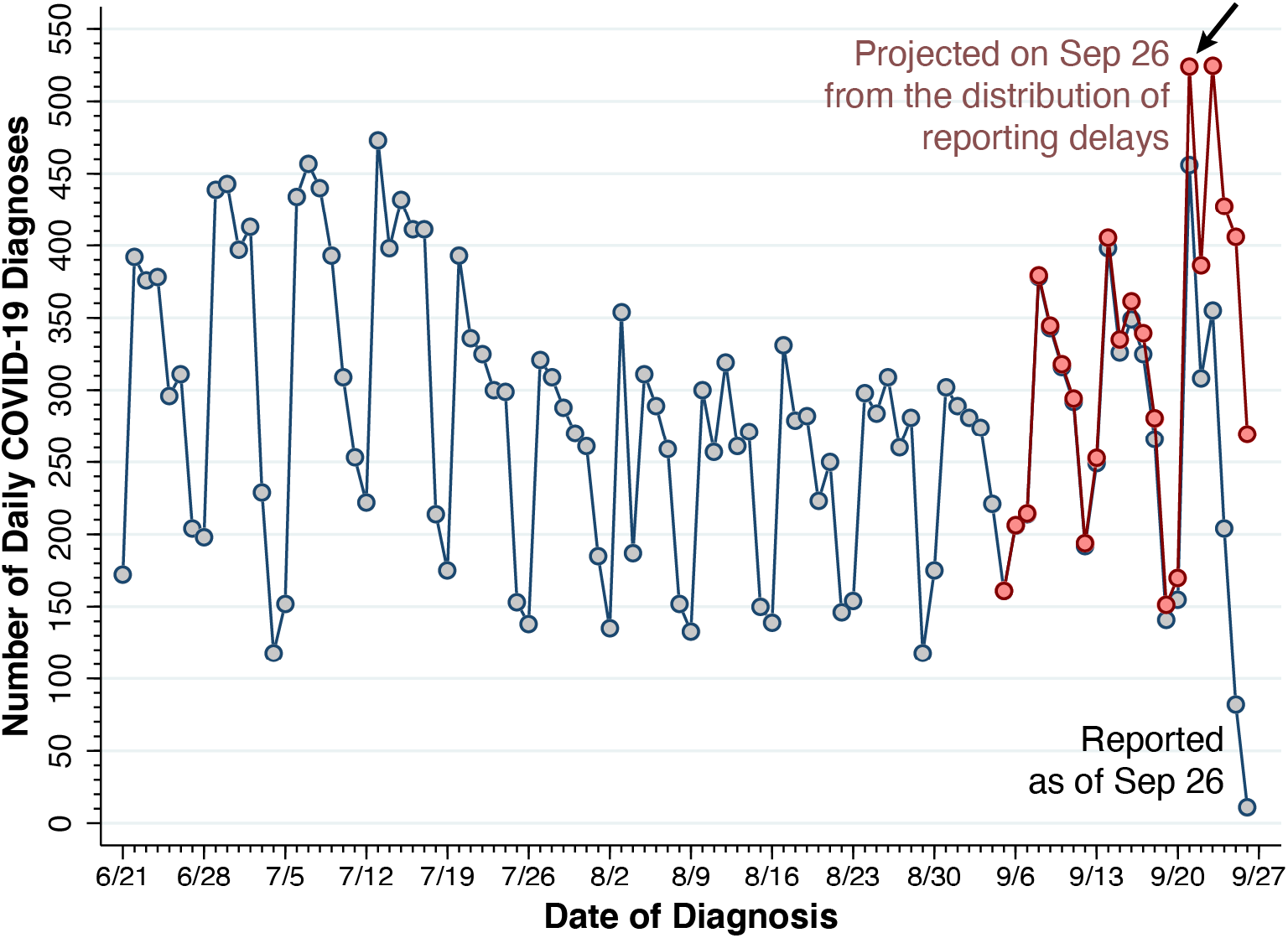
Reported and Projected COVID-19 Diagnoses, New York City, June 21 – September 26, 2020. Reported cases as of September 26 (*z*_*t*_) are indicated by gray-colored datapoints. Projected cases (*ζ*_*t*_), based up the estimated distribution 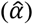 of reporting delays, are shown for the pink datapoints from September 5 – 26. The arrow at the upper right points to a projection of 524 cases for September 21, based upon 456 cases reported as of September 26 (Fig. 1) and our estimate 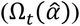 that 87.0 percent of cases had been reported by that date.

The superimposed pink datapoints the in figure, by contrast, show projected counts 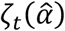 for the most recent days *t* = *T* − *n*, …, *T*, that is, from September 5 – 26. Before *t* = *T* − *n*, our projections simplify to *ζ*_*t*_ = *z*_*t*_, as reporting beyond *n* = 21 days is assumed to be complete. For September 21, as indicated by the arrow, the reported case count as of September 26 was *z*_*t*_ = 456. (See also Fig.1.) By contrast, the projected case count was 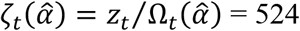, where we estimated 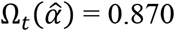 Thereafter, the projected case counts showed a continuation of the surge, reaching 524 once more on September 23, then exceeding 400 per day during September 24–25, and then displaying an expected weekend drop to 269 on Saturday, September 26.

Neither the estimated distribution of reporting delays (Fig. 2) nor the projected number of diagnosed COVID-19 cases (Fig. 3) varied significantly when we extended the observation interval backward before August 15 or increased the maximum duration of reporting delays beyond 21 days. (Results not shown.)

### Comparison of Projected and Ultimately Reported Cases

Fig. 4 compares our projected case counts (*ζ*_*t*_), again colored in pink, with case counts ultimately reported by the health department as of November 7, 2020, shown in light blue. Here, we have shifted the timeline to cover the interval from August 23 – October 11. Comparison of the two series shows significant concordance between the projected case counts and the numbers of cases ultimately reported almost 4 weeks after the end of the interval. A chi-squared test of goodness of fit failed to reject the null hypothesis that the distributions of the projected and reported case counts were equal (*χ*^2^ = 17.26, 20 degrees of freedom, yielding *p* = 0.364).

**Fig. 4.**
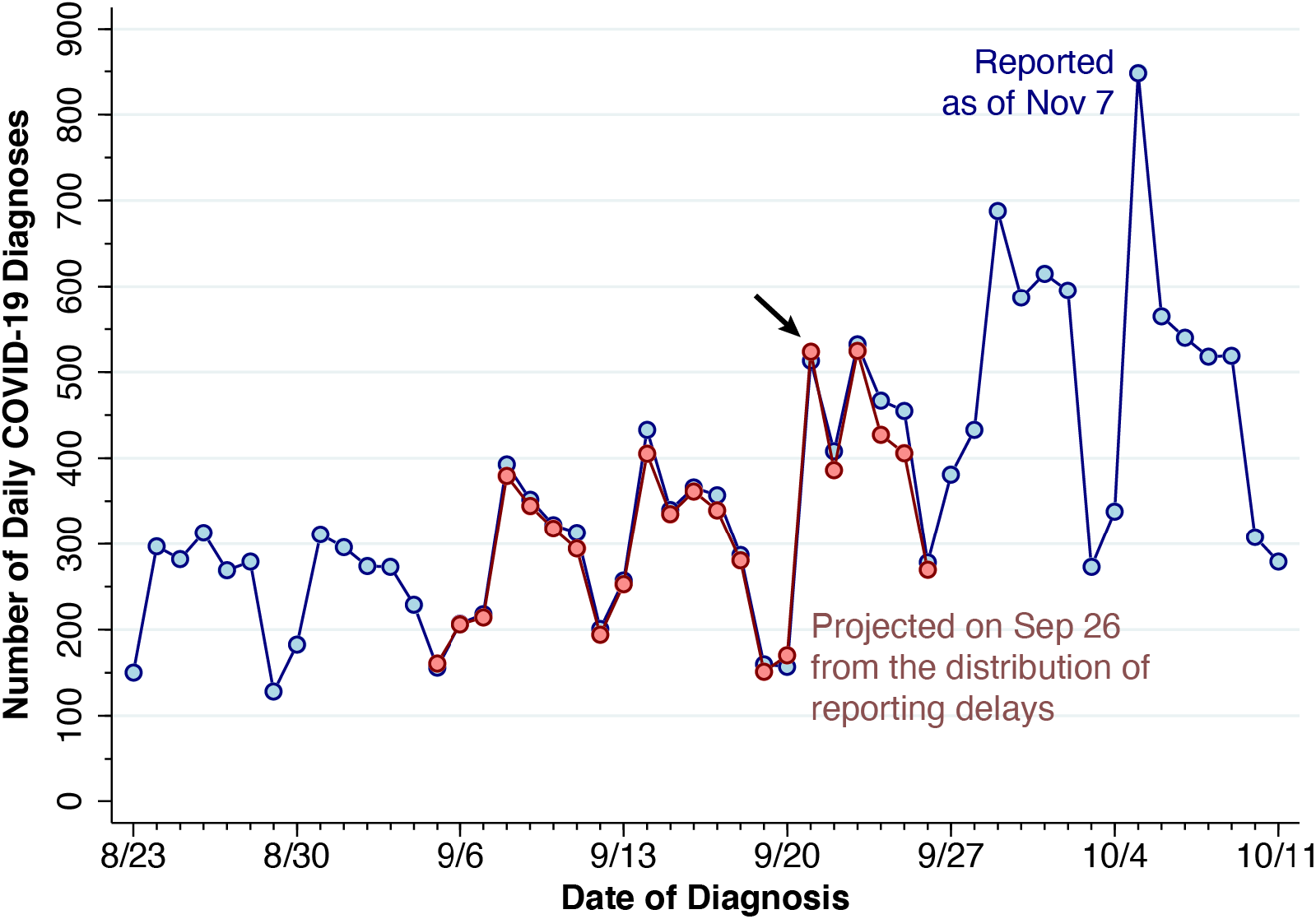
Projected and Ultimatedly Reported COVID-19 Diagnoses, New York City, August 23 – October 25, 2020. The projected case counts (Fig. 3), based upon the data available through September 26, are in pink. The ultimately reported case counts, based upon data available as of November 7, are in light blue. At the arrow, the projected count for September 21 was 524, while the ultimately reported count was 513.

Appendix Fig. B plots the projected against the ultimately reported daily case counts during September 5–26. There was no significant serial correlation of the residuals.

## Discussion

The usual workaround to address data truncation due to delayed reporting is to attach an advisory to a website graphic warning the viewer that the most recent trend is to be ignored. The key message of this article is that, so long as the distribution of reporting delays is stable, the most recently reported case counts need not be thrown out. Instead, we can use recent past data on reporting delays in order to project the population-level counts of new cases as if they had all been reported on the date of diagnostic testing.

As our study of the surge in COVID-19 cases in the fall of 2020 in New York City suggests, the resulting timely estimate of recently diagnosed cases could have had an impact on policy decisions to tighten social distancing measures. On September 29, New York City Mayor de Blasio signaled his intention to close nonessential businesses and all public and private schools in key neighborhoods of the boroughs of Queens and Brooklyn.^5^ But it was not until October 6 that New York Governor Cuomo actually intervened.^6^ As it turned out, coronavirus cases had actually been increasing throughout the city.^3^

### Diagnosis Dates Versus Report Receipt Dates

Many state and local health departments – including the New York City health department studied here – have tabulated counts of COVID-19 cases according to the date the relevant diagnostic test was *performed*. This reporting convention not only creates the data truncation problem illustrated by the precipitous drop in case counts at the far right in Fig. 1, but it also requires the public health authority to continually update past counts every time a new case report is received. These difficulties can be avoided by the alternative of reporting cases according to the date the test result was *received*. But that alternative can give a biased picture of recent incidence trends. For example, if a testing site delivers results to a public health authority in periodic batches, the reported case counts can show artifactual surges.^7^

### Absolute Case Counts Versus the Test Positivity Rate

One alternative to relying on absolute case counts is to compute positive tests as a fraction of all tests performed, an indicator often called the *test positivity rate*. This approach does not really confront the problem of reporting delays. Instead, it simply converts the problem into one of delayed reporting of test positivity rates. In fact, if negative test results are reported with a different delay distribution than positive tests, the resulting bias due to reporting delays may be exaggerated. In any event, the test positivity rate depends on the number of negative test results, which is itself an endogenous variable. Relying on a misleading decline in test positivity due to a surge in negative testing by worried well individuals, state-level officials later relaxed social distancing measures in early November when absolute case counts were still rising in the most vulnerable neighborhoods in Brooklyn.^3^

### Slow Molecular Versus Rapid Antigen Diagnostic Tests for SARS-CoV-2 Infection

Our focus here has been on *diagnostic* tests for active infection, rather than antibody tests to detect whether an individual mounted an immune response to a past infection. At the time of our study of New York City in the fall of 2020, *molecular* tests that amplify the virus’ genetic material – particularly tests based on the polymerase chain reaction (PCR) – were far and away the dominant diagnostic technology. More recently, rapid *antigen* tests of specific proteins encasing the virus’ genetic material have become widely available as an alternative.^8^ While clinics and doctors’ offices are required to report the results of these rapid antigen tests to local health agencies, individuals performing these tests at home generally do not. As a result, reported case counts are still dominated by the slower PCR technology. With molecular tests remaining the gold standard for testing,^9^ the problem of delays in publicly reported case counts of COVID-19 has not been obviated.

### Aggregate Population Delays Versus Individual-Level Delays

We have described and tested a statistical method for overcoming reporting delays at the aggregate *population level*. Our approach does not accelerate the reporting of test results at the *individual level*. We focus on the time delay from the performance of a diagnostic test to its appearance in a health department’s aggregate public tally. Our results say nothing about the time required to privately communicate test results to individual patients. Delays in informing individuals about positive and negative test results can critically influence decisions to go into or come out of isolation, and to stay away from or return to work. Technologies that facilitate communication of at-home, rapid antigen tests to public authorities may reduce delays both at the aggregate population level and the individual patient level.

### Testing Delays and Underreporting

Even with the proposed statistical correction for reporting delay, there remains the problem of *testing delay*. In the system of voluntary, symptom-motivated testing in the United States, testing delay has two components. The first is the incubation period between initial infection and first symptoms of illness, initially estimated to be about 5 days for the ancestral strain of SARS-CoV-2, about 4 days for the Delta variant, and closer to 3 days for the more recent Omicron variant.^10^ The second is the additional delay between the onset of symptoms and date the test is performed. Widely available rapid antigen testing may have partially reduce the second bottleneck.

Quite apart from the issue of testing delay, there is now growing evidence that during the Omicron wave, as many as 75 percent of all SARS-CoV-2 infections have not been reported by public authorities.^11^ While some of this underreporting has been the result of the growing use of in-home rapid antigen tests, there has been a surge in asymptomatic and mildly symptomatic infections. Quantitative modeling of underreporting remains a challenging problem.

### Technical Issues

The statistical method proposed here assumes that reports arrive according to an independent, homogeneous Poisson process. Reporting delays could have varied according to laboratory of diagnosis, duration or severity of infection, and characteristics of the individual patient. As already noted, reports could also have arrived in batches. Data to test these possibilities were unavailable.

## Data Availability

Data, programs and output are posted on the Open Science Framework at https://osf.io/u4svy/.

## Appendix

Fig. A compares the estimated cumulative distribution of reporting delays for two non-overlapping time periods: June 21 – August 1; and August 15 – September 26, 2020. Each curve plots the estimated cumulative percentage of diagnosed cases reported up to and including the delay interval measured on the horizontal axis. With 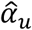 denoting the estimated probability that a diagnosed case will be reported with delay *u*, the figure thus plots 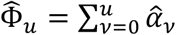 as a function of the reporting delay time *u*. Comparison of the two cumulative distribution functions shows a significant reduction in the duration of reporting delays during the summer and early fall of 2020. As noted in the figure, the respective estimates for 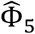 were: 0.652 for the earlier interval from June 21 – August 1; and 0.870 for the later interval from August 15 – September 26.

**Fig. A.**
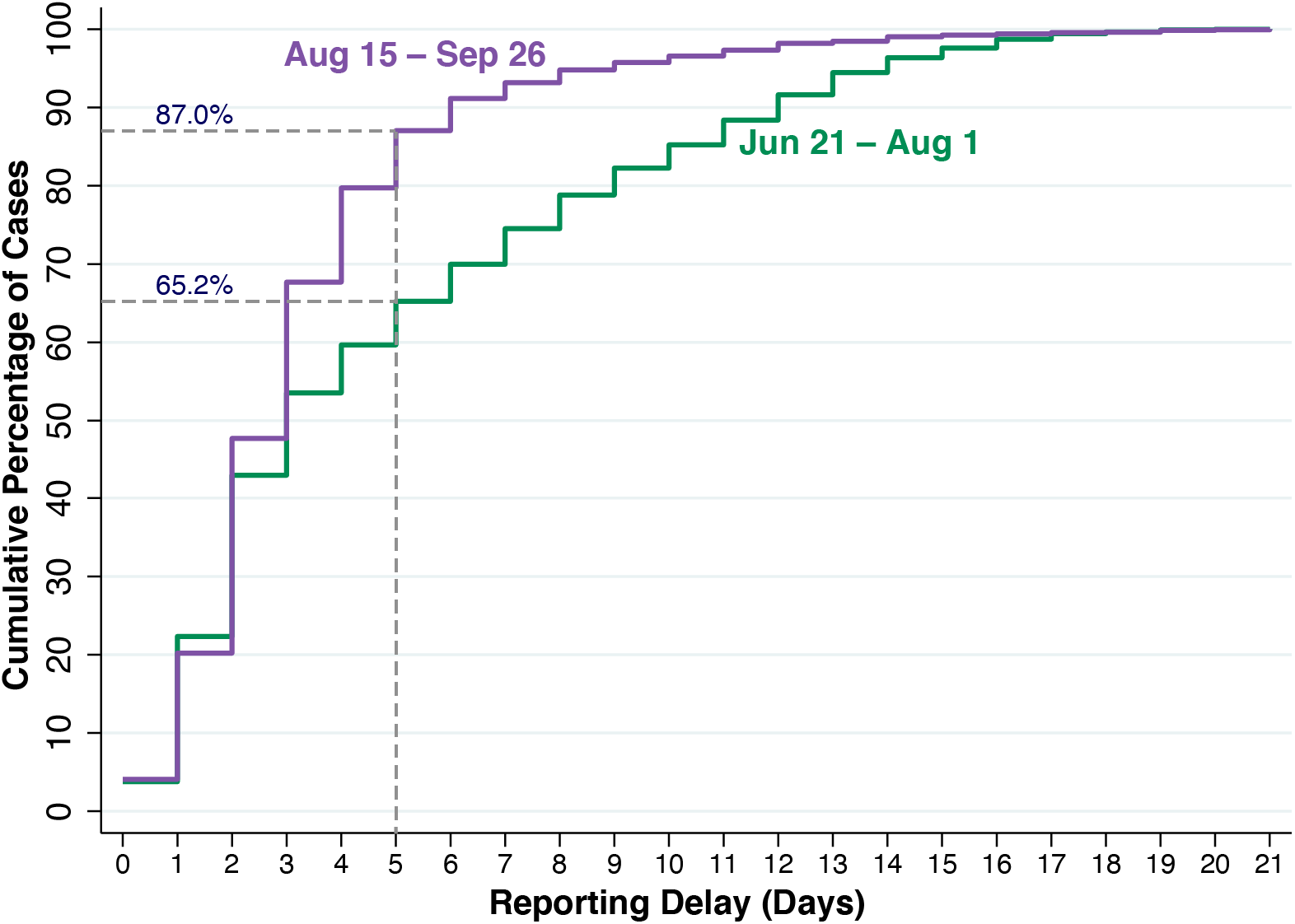
Estimated Cumulative Distribution of COVID-19 Reporting Delays, New York City, June 21 – August 1 (Green) and August 15 – September 26, 2020 (Purple). During June 21 – August 1, an estimated 65.2% of cases were reported within 5 days of the date when the diagnostic test was performed. During August 15 – September 26, this proportion had increased to 87.0%. The mean reporting delay was 4.96 days during June 21 – August 1 and 3.31 days during August 2 – October 2.

Fig. B plots the projected daily case counts for September 5–26, based upon the data available as of September 26, against the reported daily case counts for the same interval, based upon the data ultimately made available on November 7. Superimposed is a 45-degree line indicating equality between the two variables. The serial correlation coefficient of the projected-versus-reported residuals was 0.266. We could not reject the null hypothesis of serially uncorrelated residuals (*p* = 0.244).

**Fig. B.**
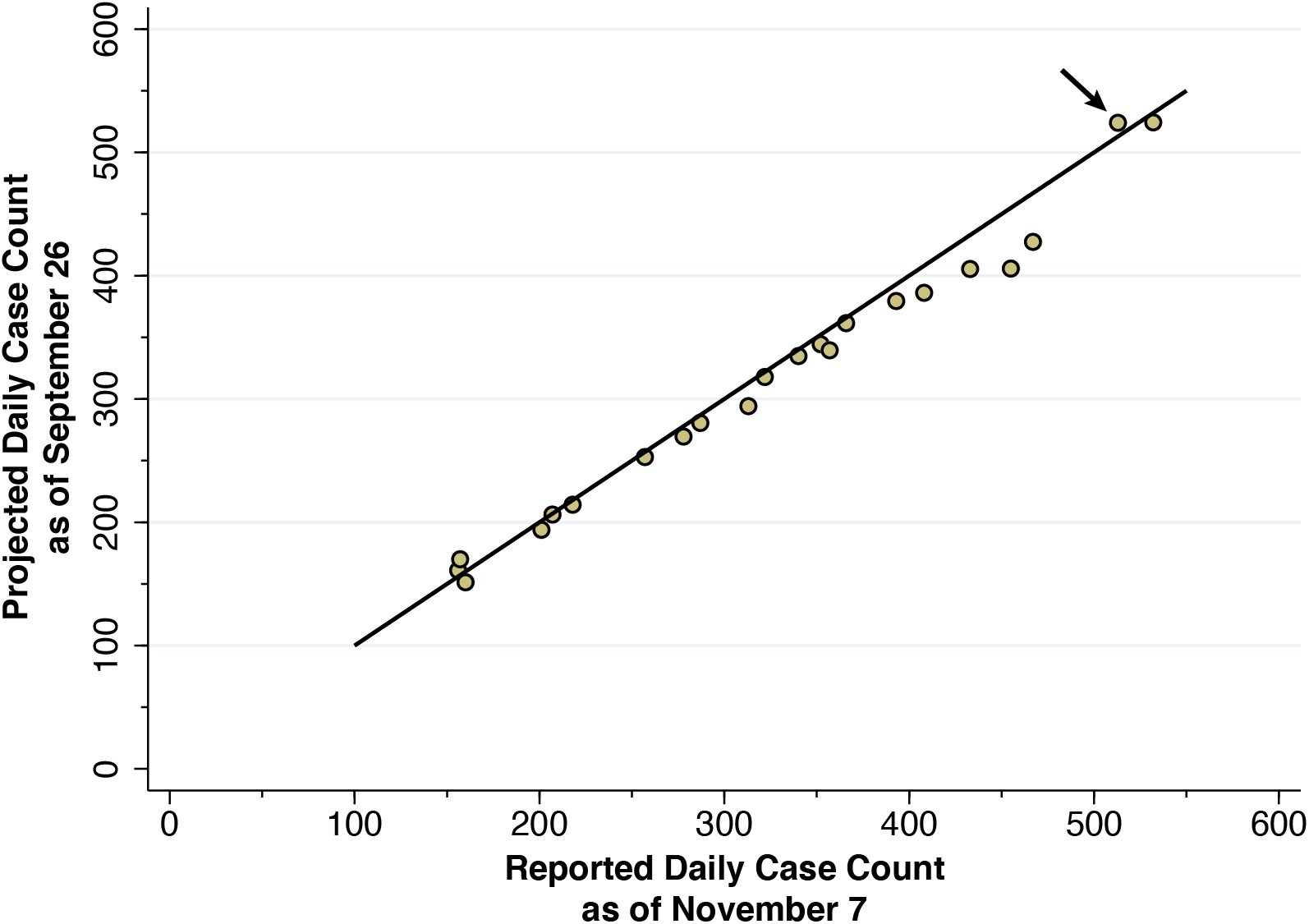
Projected Daily Case Count as of September 26 Versus Reported Daily Case Count as of November 7. The superimposed 45-degree line indicates equality between the two variables. The arrow shows the data for September 21, where the projected count was 524 and the ultimately reported count was 513.

